# Depression, Anxiety and Depression-anxiety comorbidity amid COVID-19 Pandemic: An online survey conducted during lockdown in Nepal

**DOI:** 10.1101/2020.04.30.20086926

**Authors:** Anil Sigdel, Anu Bista, Navaraj Bhattarai, Bimal Chandra Pun, Govind Giri, Hannah Marqusee, Subash Thapa

## Abstract

**Background:** Little is known about the effect of the COVID-19 pandemic on mental health status during the lockdown period. Therefore, this study was conducted to assess prevalence of depression, anxiety and depression-anxiety comorbidity, and associated factors during the COVID-19 lockdown in Nepal.

**Methods:** A quantitative cross-sectional study was conducted among the general population of Nepal. Data was collected from April 9 to April 16, 2020 using an e-questionnaire which was shared through different popular social media. A total of 349 participants were included. Self-reported depression and anxiety were assessed using the Patient Health Questionnaire and Generalized Anxiety tools respectively. Logistic regression analysis was conducted to identify the factors associated with depression, anxiety and depression and anxiety co-morbidity.

**Results:** The prevalence rates of depression, anxiety and depression-anxiety co-morbidity were found to be 34.0%, 31.0% and 23.2% respectively. The multi-variate analysis showed that females, those living alone, health professionals and those who spent more time in accessing information about COVID-19 were significantly more likely to have depression, anxiety and depression-anxiety co-morbidity.

**Conclusions:** High rates of depression and anxiety and co-morbidity were found to be prevailing among the general population during the COVID-19 pandemic lockdown in Nepal. The results suggest that only the scientific, but contextually appropriate messages about the disease should be disseminated to reduce unnecessary fears and anxiety. Awareness interventions to promote mental wellbeing need to be integrated into the response interventions. Community mental health care should be made accessible to at-risk groups.

## Introduction

A novel coronavirus, SARS-CoV-2, which was first noted in the Wuhan city of China on December 2019, is rapidly spreading globally (1). The World Health Organization (WHO) has declared this disease as a Public Health Emergency of International Concern (PHEIC) on 30 January 2020, which is the sixth time WHO has declared a PHEIC after the International Health Regulations (IHR) that came into effect in 2005. Thereafter has been a rapid increase not only in infections and deaths, but also anxieties, stigma, mistrust, and rumor-mongering among the public (2). WHO has reported a total of 2,436,743 confirmed cases and 165,310 deaths across 206 countries/territories by April 22, 2020 (3). The COVID-19 pandemic is on its way to cause historically significant global change (4), as it continues to rise. The World is on high alert; borders are closed and strict measures are being taken to control the spread of COVID-19 (5). At the same time, many countries affected by COVID-19 have implemented temporary lockdowns restricting people’s unnecessary movement outside the home and ensuring that people stay safe at home.

Multiple biological and behavioral pathways are likely contributing to the linkages between mental health conditions and viral diseases, such as COVID 19 (6). Individuals or communities experience a mental instability along with social and economic losses which might precipitate as mental stress, anxiety and depression (7). It is reported that nearly all people affected by or during such global emergencies will experience some level of psychological distress, which for most will improve over time. The prevalence of common mental disorders are expected to be more than double (8). For instance, the estimated prevalence of mental disorders among the conflict-affected population according to WHO is 13% for mild forms of depression, anxiety, and post-traumatic stress disorder and 4% for moderate forms of these disorders. Depression and anxiety are found to increase with age and are more common in women than in men in a conflict-affected setting (8). The burden of mental problems would be even higher due to the fact that healthcare, and particularly the mental health care system, is severely affected due to COVID-19.

Several scholars have provided varied propositions why emergency situations, such as pandemics are associated with the rise of mental and emotional problems. As suggested by Nilamadhav Kar, based on a study conducted after a terrorist initiated bomb blast incident in India in 1996, people go through different negative mental and emotional states, including helplessness, severe stress, severe mood swings and forgetfulness, emotional instability, anxiety, stress reactions, and trauma (9). On the contrary, Wachinger observed such mental changes, during an emergency situation, domain as a protective factor (a coping mechanism) (10) and stated that such changes included willingness to control the emotional extremes, self-regulation of one’s emotions, inculcating hope and courage, positive attitude and acceptance of the situation, concern about oneself and family members and ability of the individual to prepare oneself, which could in turn solidify the emotional instability (7). Among the variety of mental and emotional changes that occur during such situations, some of them more strongly improve the general wellbeing of individuals, but anxiety and depression are among the most common problems (global prevalence is estimated to be between 3.6% to 4.4%) (11).

Anxiety and depression as medical problems include generalized and acute anxiety, post-traumatic stress disorder (PTSD), phobias, panic disorder, major depressive disorder, bipolar illness, and other mood disorders. (11) There are known effective treatments for such mental disorders, but unfortunately between 76% to 85% of people from low and middle-income countries receive no treatment for their mental illness (12). A study done among physicians in China (the most affected country by COVID19 in early days) has shown that an estimated 25.67% of physicians had anxiety symptoms, 28.13% had depressive symptoms, and 19.01% had both anxiety and depressive symptoms. Poor self-reported physical health, frequent workplace violence, lengthy working hours (more than 60 hours a week), frequent night shifts (twice or more per week), and lack of regular physical exercise were found to be associated with anxiety and depression symptoms among physicians (13). There are various other factors, for instance the displacement of the family, death of a loved one, socio-economic loss, environmental loss, lack of mental preparedness for disaster, lack of social support and negative coping skills, that might lead to the psychological vulnerabilities of those affected (9). Several studies have evaluated the psychological status of people during pre and post emergency, however there are limited studies that assess the mental suffering of people during the course of epidemics. As such, there is an unmet need for greater understanding of the management of anxiety and depression in any epidemic or pandemic situation (14). In any pandemic, the most vulnerable communities are in developing-world cities where there are huge numbers of people, with poor health systems and where millions lack access to services (14).

According to Health Emergency Operation Center (HEOC), the first case in Nepal was reported on January 13, 2020 and to date the situation is in control with only 45 reported cases (7 recovered, 38 in isolation) (15). With the second case, Nepal announced a complete lockdown from 24 March 2020 as a measure to control community transmission of COVID-19. The vulnerability of Nepal increases with its open border with India (with 20,178 reported cases and 645 deaths) and China (being the epicenter for the COVID-19 with 82,788 cases and 4,632 deaths) as of April 22, 2020 (3). On one hand, the development of social media and social networking has made it easier to share the severity and extent of the damages while on the other hand it has increased the higher levels of indirect exposure, leading to increased risk for stress, anxiety, and depression from indirect trauma (16). The bombarding of breaking news from the media is also creating havoc on people’s mental status. Although the priority is to stop the transmission, provide care to those infected, and to seek a treatment and vaccine for long-term management, management of mental health problems has not received any attention and therefore, there is a need to investigate and explain the increased burden of mental health problems and the factors associated with it. The present study is aimed at generating evidence on the prevalence of anxiety and depression among the general population and the factors associated with anxiety and depression during the COVID-19 pandemic lockdown in Nepal.

## Materials and Methods

### Study Design and Study Population

This was a cross-sectional survey study to assess the prevalence and factors associated with depression, anxiety and depression-anxiety comorbidity among the general population of Nepal. The participants of this study were a sample of the general population with access to internet; who were aged 18 years old and above, and who were able to provide written consent.

### Sampling and recruitment

As the Government of Nepal recommended the public to minimize face-to-face interactions and isolate themselves at home, snowball sampling method was used to enroll the participants for the survey. Advertisements for the survey were shown to the general people on popular social platforms: Ministry of Health and Population (MoHP) Facebook page, Nepal Public Health Association (NEPHA) Facebook page, Pokhara University Facebook page and Purbanchal University Facebook page. A total of 355 participants participated in the study, where only 350 (98.6%) provided consent for the study, and one participant was younger than 18 years and thus was excluded from the study. Hence, the total sample included in the study was 349.

### Questionnaire and Measurement

A self-administrated structured questionnaire was developed. The questionnaire was grouped into four sections: (a) socio-demographic section; (b) exposure to media; (c) Anxiety and (d) Depression. The questions related to socio-demography and media exposure were adopted from the Nepal Health and Demographic Survey (NDHS) 2016 (17). Generalized Anxiety Assessment (GAD-7), a self-administered questionnaire having sensitivity of 89% and specificity of 82%, developed by Robert L. Spitzer et al., was used as a tool to measure the anxiety among participants (17). Likewise, the Patient Health Questionnaire (PHQ-9) is a self-administrated questionnaire developed by Drs. Robert L. Spitzer et al. which was used to measure the depression among participants (18). The PHQ-9 tool has a sensitivity of 88% and specificity of 88% for major depression (18).

*Outcome variables:* The outcome variables are outlined in **Table *1***. Anxiety and depression were the outcome variables for this study. Anxiety was ascertained using the GAD-7 tool. GAD-7 consists of a 7 items questionnaire that asked participants how often, during the last 2 weeks, they were bothered by each symptom. Response options were “not at all,” “several days,” “more than half the days,” and “nearly every day,” scored as 0, 1, 2, and 3 respectively (17). The sum of all the items of GAD-7 were used to measure the level of anxiety. A score of up to 5 was considered mild; 6-10 was considered moderate; 11-15 was considered moderately severe anxiety and 15-21 was considered severe anxiety. The score was dichotomized for logistic regression as the sum of GAD-7 less than 10 was considered normal and a GAD-7 score greater than or equal to 10 were considered as participants with anxiety.

**Table 1.** Outcome and independent variables of the study

| SN | Variables | Categories of Variables |
| --- | --- | --- |
| **Outcome Variables** |  |  |
| 1 | Anxiety among general population | Normal (score of 10 or less) and Anxiety (score of more than 10) based on Generalized Anxiety Assessment. (GAD-7) |
| 2 | Depression among general population | Normal (score of 10 or less) and Depression (score of more than 10) based on Patient Health Questionnaire (PHQ-9) |
| 3 | Comorbidity of depression and anxiety | Yes (participants with both depression and anxiety); No |
| **Independent Variables** |  |  |
| **Socio-demographic Variables** |  |  |
| 1 | Sex of Participants | Male; Female |
| 2 | Age of Participants | Less than 20 years; 20-29 years; 30-39 years; and 40 years and above |
| 3 | Place of current residence | Urban (Metropolitan/sub-metropolitan); Semi-Urban (Municipalities) and Rural (Rural Municipalities) |
| 4 | Ethnicity | Brahmin/Chhetri; Janajati/Adabasi and Others. Adopted from Health Management Information System (HMIS) of Government of Nepal |
| 5 | Educational level | Primary level (Up to grade 5); Secondary level (grade 6-10); Higher Secondary level (grade 11-12) and Graduate and above |
| 6 | Religion | Hindu; Buddhist; Muslim; Kirat; Christian; Others. Adopted from NDHS 2016 |
| 7 | Marital Status | Not in married/not in union; Married; Divorced/separated; Widow/Widower. Adopted from NHDS 2016 |
| 8 | Major Occupations | Job Holders (Formally employed); Students; Businessperson; Labor/Daily Wages; Agriculture; Others |
| 9 | Household ownership | Own house; Rented House |
| 10 | Currently accompanying status | Alone; Family |
| 11 | Health Professional | Yes; No |
| **Media Exposure** |  |  |
| 1 | Most used mass media to get information on COVID 19 | Television; Radio; Social Media; Online Newspaper; Print Newspaper; Friends; Others |
| 2 | Average hours spent on mass media on a day | Time in hours. |

To ascertain depression among the participants, the PHQ-9 questionnaire was used. The PHQ-9 consists of nine questions that asked participants how often, during the last 2 weeks, they were bothered by each symptom. Response options were “not at all,” “several days,” “more than half the days,” and “nearly every day,” scored as 0, 1, 2, and 3 respectively. The sum of the scores of all nine items of the PHQ-9 were used to determine the level of depression. A score of up to 5 was considered mild; 6-10 was considered moderate; 11-15 was considered as moderately severe depression and 16 and above were considered as severe depression (19). However, for logistic regression, the score was dichotomized as participants with depression (the PHQ-9 score of more than 10) and normal (the PHQ-9 score of 10 and less).

*Independent variables:* Independent variables such as socio-demographic factors (sex, age, place of current residence, caste/ethnicity, educational level, religion, marital status, major occupation, accompanying status and household ownership), and Media Exposure (most used mass media to get information on COVID-19 and average time spent on mass media in a day) were collected from the participants.

## Data collection

A survey form was developed using a google form and participants for the survey were electronically invited through personal email and Facebook messenger. Data collection for the survey was conducted within the first week of the lockdown in Nepal, i.e. from April 9, 2020 to April 16, 2020. The survey form was set up in such a way that one participant can only submit one form with one google account.

## Ethical Considerations

Ethical approval was obtained from Nepal Health Research Council (NHRC), Kathmandu. The objectives, risks, benefits and use of study was shared with each participant in written form prior to administration of the questionnaire. The system was set up in such a way that participants could only answer the questionnaire once consent was provided. Written informed consent was obtained from participants prior to administration of questions. No personal identifiers were collected during the interview and no personal identifiers were disclosed anywhere in the study.

## Data Analysis

Data were analyzed using Statistical Software Social Sciences (SPSS) version 20 for windows. Descriptive statistics were calculated for socio-demographic variables, media exposure, anxiety and depression. A chi-square test was applied to test the statistical significance among outcome and independent variables. Binary logistic regression analysis was carried out to identify factors associated with anxiety and depression. All the variables with a p-value of 0.20 in bivariate analysis were entered in a multivariate analysis model. Forward stepwise logistic model was used with a p value of 0.20 for entry and 0.10 for exit. Hosmer and Lemeshow goodness of fit was used to test the fitness of model and Variance Inflation Factors (VIF) was used to measure the multi-collinearity among independent variables. All the predicator variables have VIF less than 2.

## Results

More than half (54.2%) of participants were male and the mean age of participants was 27.8 years. Nearly two-third of the participants (62.5%) belong to Brahmin or Chettri ethnic group followed by Janajati/Adhibasi (26.5%). A majority of the participants (91.1%) had completed Bachelor level and were Hindu by religion (90.5%) Nearly two-thirds (62,2%) were single. A majority were job holders (57%), reside in urban areas (65%), were living with family (72.5%), living in their own house (58.2%) and were health professionals (60%) as summarized in Table 2.

**Table 2.**
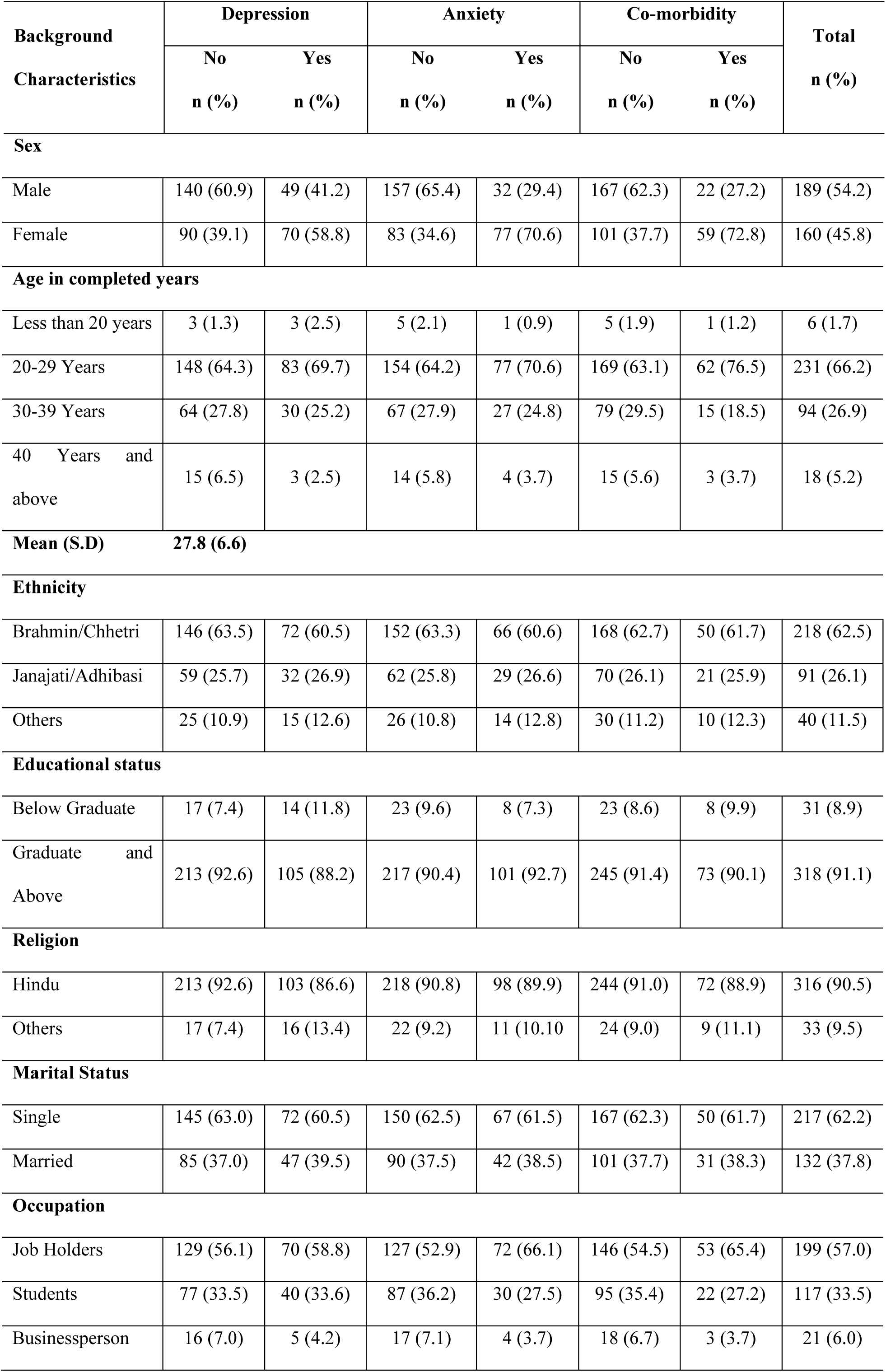
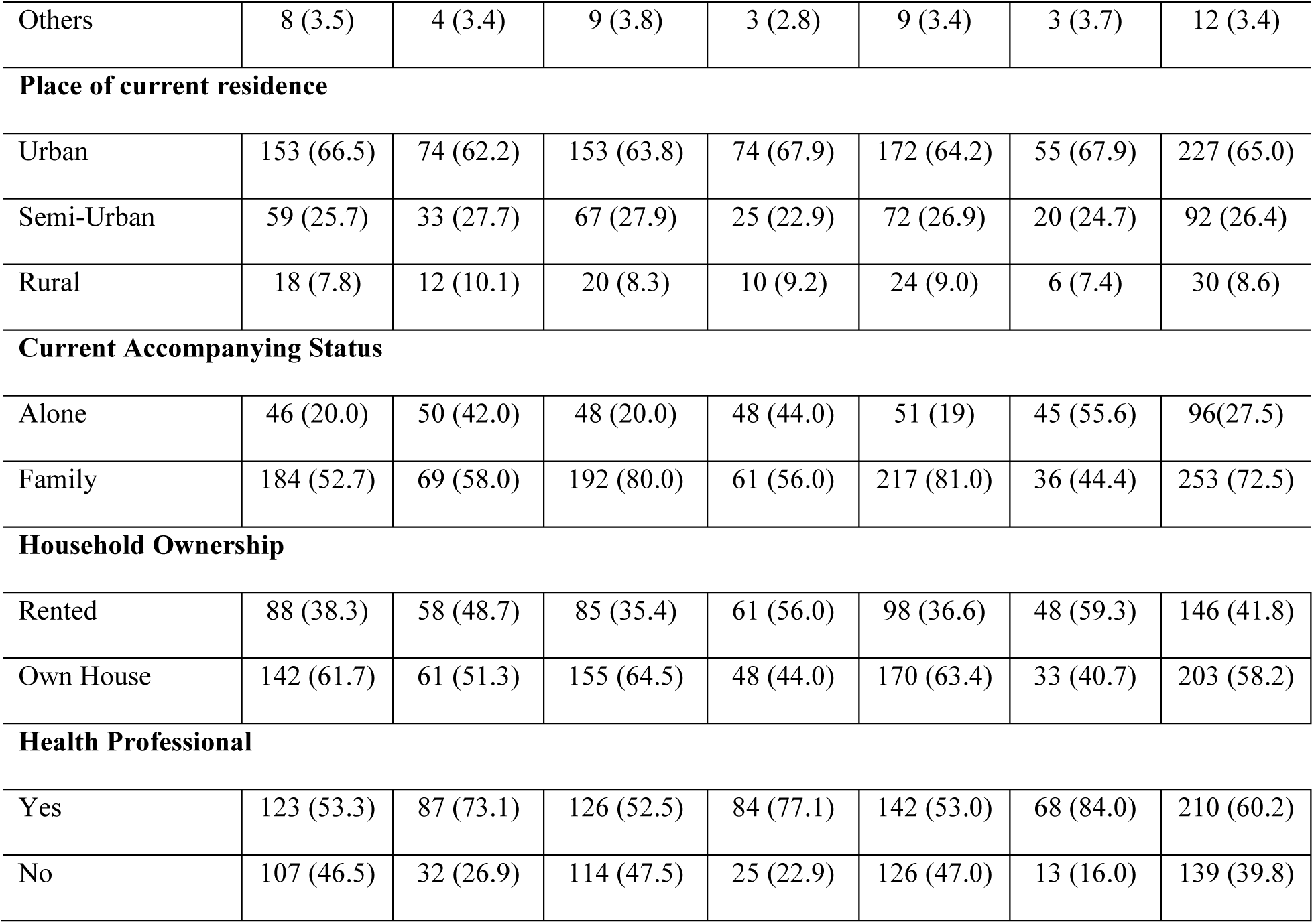
Socio-demographic characteristics of the study participants

More than two-thirds (69.3%) used social media for accessing information on COVID-19 followed by online newspapers (14.6%) and the mean time spent accessing information on COVID-19 was 3.4 hours per day as summarized in **Table *3***.

**Table 3.**
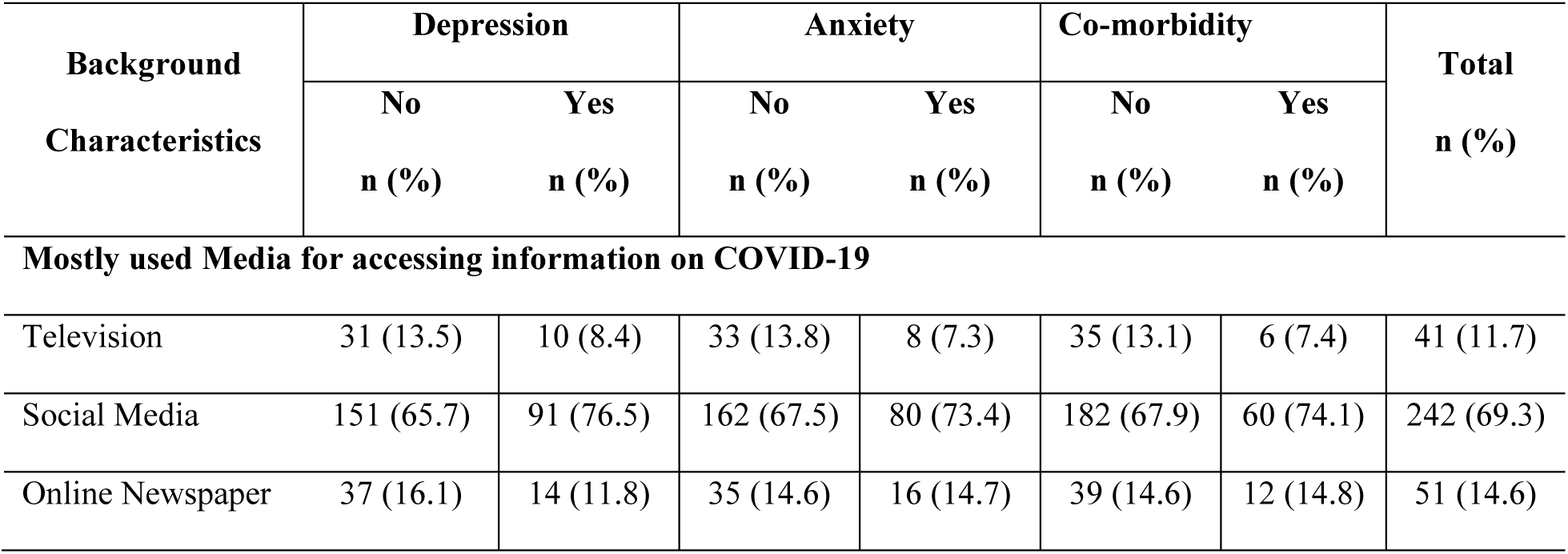
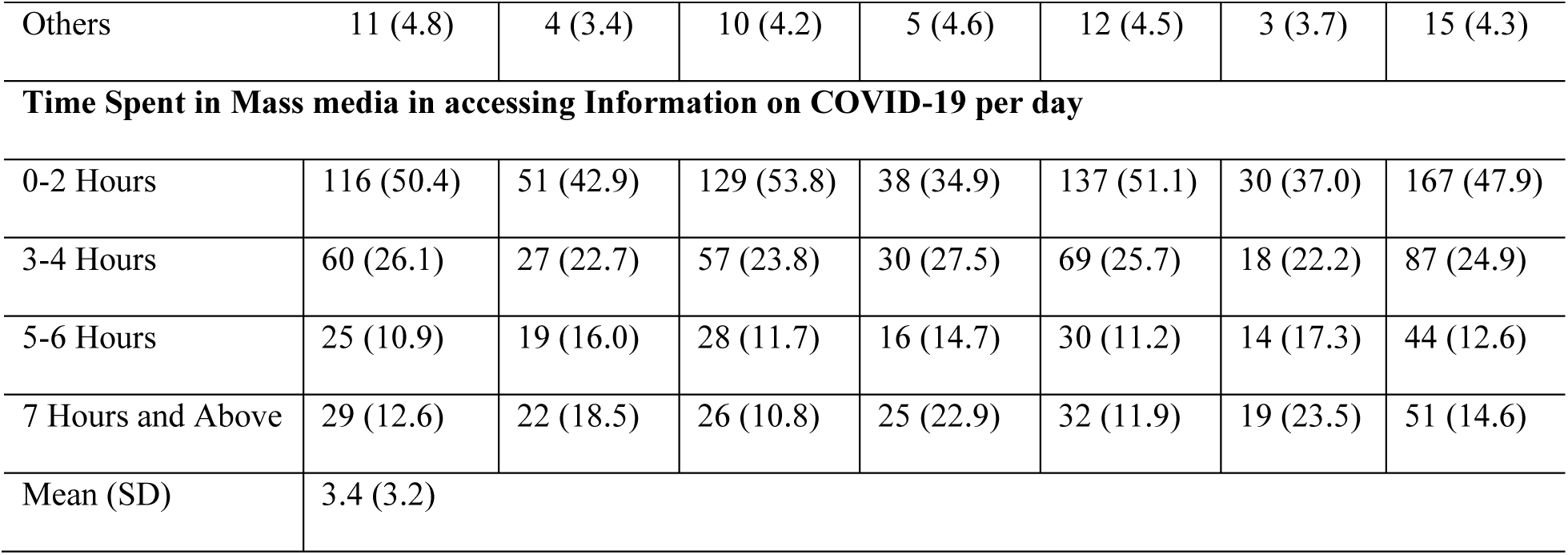
Media Exposure on accessing information on COVID-19

| Background<br>Characteristics | Depression |  | Anxiety |  | Co-morbidity |  | Total<br>n (%) |
| --- | --- | --- | --- | --- | --- | --- | --- |
|  | No | Yes | No | Yes | No | Yes |  |
|  | n (%) | n (%) | n (%) | n (%) | n (%) | n (%) |  |
| Mostly used Media for accessing information on COVID-19 |  |  |  |  |  |  |  |
| Television | 31 (13.5) | 10 (8.4) | 33 (13.8) | 8 (7.3) | 35 (13.1) | 6 (7.4) | 41 (11.7) |
| Social Media | 151 (65.7) | 91 (76.5) | 162 (67.5) | 80 (73.4) | 182 (67.9) | 60 (74.1) | 242 (69.3) |
| Online Newspaper | 37 (16.1) | 14 (11.8) | 35 (14.6) | 16 (14.7) | 39 (14.6) | 12 (14.8) | 51 (14.6) |
| Others | 11 (4.8) | 4 (3.4) | 10 (4.2) | 5 (4.6) | 12 (4.5) | 3 (3.7) | 15 (4.3) |
| **Time Spent in Mass media in accessing Information on COVID-19 per day** |  |  |  |  |  |  |  |
| 0-2 Hours | 116 (50.4) | 51 (42.9) | 129 (53.8) | 38 (34.9) | 137 (51.1) | 30 (37.0) | 167 (47.9) |
| 3-4 Hours | 60 (26.1) | 27 (22.7) | 57 (23.8) | 30 (27.5) | 69 (25.7) | 18 (22.2) | 87 (24.9) |
| 5-6 Hours | 25 (10.9) | 19 (16.0) | 28 (11.7) | 16 (14.7) | 30 (11.2) | 14 (17.3) | 44 (12.6) |
| 7 Hours and Above | 29 (12.6) | 22 (18.5) | 26 (10.8) | 25 (22.9) | 32 (11.9) | 19 (23.5) | 51 (14.6) |
| Mean (SD) | 3.4 (3.2) |  |  |  |  |  |  |

The overall prevalence rates of depression, anxiety and depression and anxiety co-morbidity were found to be 34.1% (95% CI: 28.8-39.5), 31.2% (95% CI: 26.4-36.4) and 23.2% (95% CI: 18.9-27.5) respectively (see **Table *4***). The prevalence rates of depression, anxiety and depression-anxiety comorbidity were found to be higher among female participants than male participants (as see Table 1). ***Fig 1 shows*** Box and Whisker plot for the mean (depression:0; anxiety: 0), median (depression: 8, anxiety: 7), quarter 1 (depression: 3, anxiety: 3), quarter 3 (depression: 14, anxiety: 12) and interquartile range (depression: 11, anxiety: 9) of depression, anxiety and depression-226 anxiety co-morbidity.**Fig *1***

**Table 4.** Prevalence of depression, anxiety and co-morbidity of anxiety and depression

| Prevalence of depression | n (%) | 95% CI |
| --- | --- | --- |
| Normal | 230 (65.9) | 60.5-71.1 |
| Depression | 119 (34.1) | 28.9-39.5 |
| Prevalence of anxiety |  |  |
| Normal | 240 (68.8) | 63.6-73.6 |
| Anxiety | 109 (31.2) | 26.4-36.4 |
| Prevalence of depression-anxiety co-morbidity |  |  |
| Normal | 268 (76.8) | 72.5-81.1 |
| Co-morbidity | 81 (23.2) | 18.9-27.5 |

**Fig 1.**
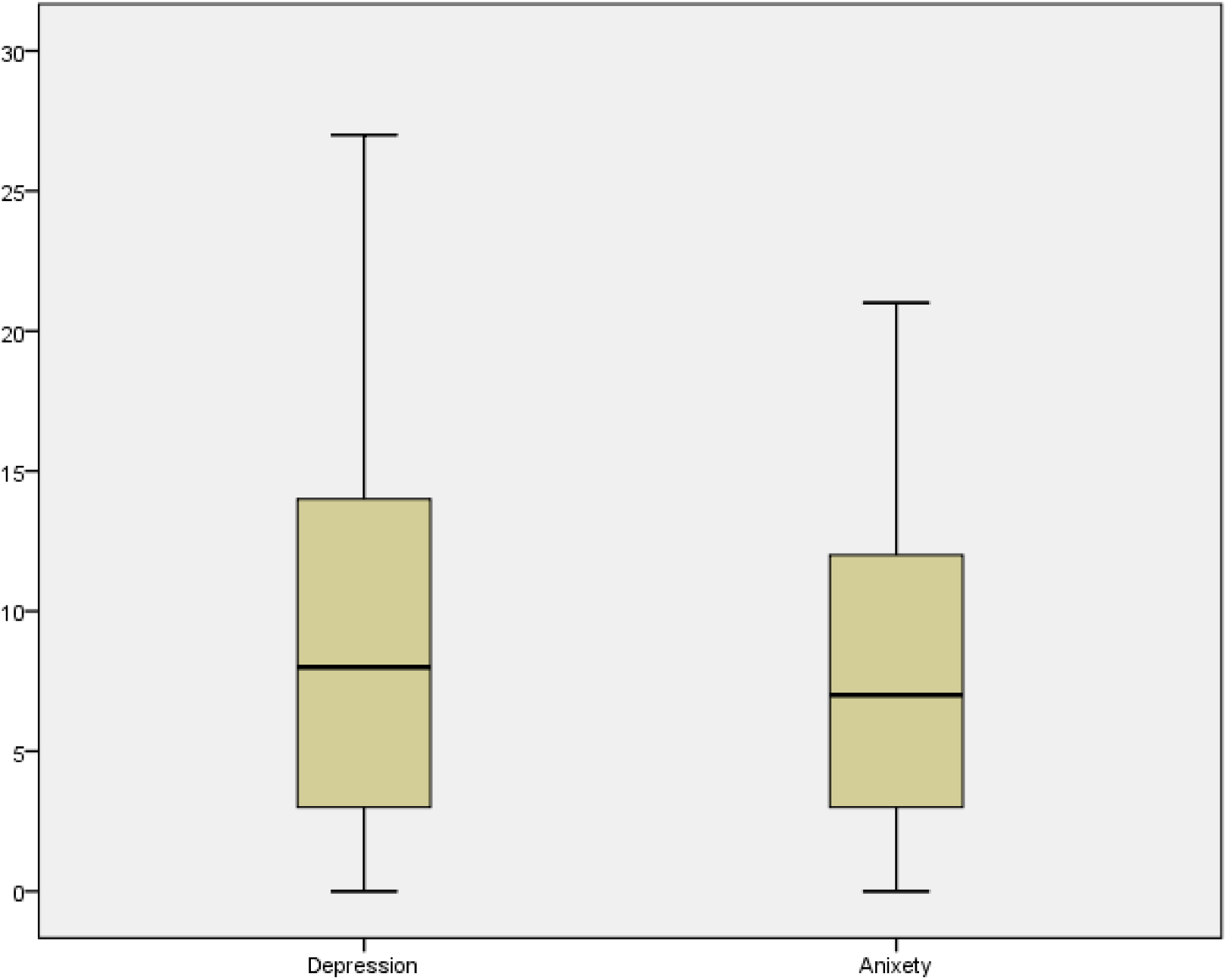
Box and Whisker plot

Forward stepwise logistic regression was carried out with depression and co-morbidity of depression and anxiety as outcome variables and five predictor variables with a p value less than 0.2. Logistic regression was also carried out with anxiety as an outcome variable and six predicator variables with a p value less than 0.20 in bivariate analysis. The final model is summarized in ***Table 5***. Female participants were 2.4 times more likely to be depressed (95% CI: 1.48-3.89); 6.3 times more likely to be anxious (95% CI: 3.54-11.18) and 7.4 times more likely to have the co-morbid condition. Those participants who were currently living alone were found to be more depressed (AOR: 3.5; 95% CI: 1.82-6.84), anxious (AOR: 3.3; 95% CI: 1.65-6.74) and have the co-morbid condition (AOR: 8.8, 95% CI: 3.85-20.12) compared to those who were accompanied by their family. Likewise, health professionals compared to others had 1.7 times, 2 times and 3.4 times higher odds of being depressed (95% CI: 1.04-2.90), anxious (95% CI: 1.16-3.69) and having co-morbid condition (95% CI: 1.62-6.97) respectively. Also, time spent in mass media in accessing COVID-19 information was found to be significantly associated with depression (AOR: 1.1, 95% CI: 1.01-1.16), anxiety (AOR: 1.1, 95% CI: 1.08-1.27) and depression and anxiety co-morbidity (AOR: 1.2, 95% CI: 1.06-1.26). However, place of current residence of participants and household ownership were not found to be significantly associated with depression, anxiety and depression and anxiety comorbidity.

**Table 5.** Factors associated with depression, anxiety and depression-anxiety co-morbidity

| Background Variables | Depression<br>AOR (95% CI) | Anxiety<br>AOR (95% CI) | Co-morbidity<br>AOR (95% CI) |
| --- | --- | --- | --- |
| **Sex** |  |  |  |
| Male | 1 | 1 | 1 |
| Female | 2.4 (1.48-3.89) | 6.3(3.54-11.18) | 7.4 (3.73-14.81) |
| **Currently Accompanying Status** |  |  |  |
| Family | 1 | 1 | 1 |
| Alone | 3.5 (1.82-6.84) | 3.3 (1.65-6.74) | 8.8 (3.85-20.12) |
| **Health Professionals** |  |  |  |
| No | 1 | 1 | 1 |
| Yes | 1.7 (1.04-2.90) | 2.0 (1.16-3.69) | 3.4 (1.62-6.97) |
| **Household Ownership** |  |  |  |
| Rented House | 1 | 1 | 1 |
| Own House | 1.4 (0.80-2.72) | 0.8 (0.44-1.62) | 1.38 (0.64-2.98) |
| **Hours spent in mass media in accessing COVID-19 information** | 1.1 (1.01-1.16) | 1.1 (1.08-1.27) | 1.2 (1.06-1.26) |
| **Place of Current Residence** |  |  |  |
| Rural |  | 1 | 1 |
| Urban |  | 0.8 (0.31-2.15) | 1.5 (0.48-5.02) |
| Semi-Urban |  | 0.6 (.23-1.88) | 1.3 (0.37-4.51) |

AOR= Adjusted Odds Ratio; CI= Confidence Interval; Reference Category=1

## Discussions

Along with socio-economic distress, psychological distress also occurs simultaneously in the victims of emergency. Depression and anxiety, the most common mental disorders are assumed to increase as an outcome of negative impact on mental health (9). The lockdown policies in response to COVID19 have been understood as the world’s biggest psychological experiment. For instance, a study conducted in China after the declaration of COVID-19 emergency situation clearly relates the theory of Behavioral Immune System (BIS), as most people showed negative emotions (such as anxiety, depression, and indignation) and less people showed positive emotions (Oxford happiness) which means more people did produce negative emotions for their self-protection (20,21). These results are consistent with previous studies as well, which found that public health emergencies (e.g., SARS) and the circumstances created by such emergencies, such as forced quarantine and lockdown, may generate a series of stress emotional responses including higher levels of anxiety and other negative emotions (22,23).

A study done in Indonesia shows that the prevalence rates of PTSD, depression, and anxiety were 58.3%, 16.8% and 32.1%, respectively following the earthquake in 2016(24). Another study done in China after the declaration of COVID-19 as a public health emergency of international concern (PHEIC) by WHO shows that 53.8% of respondents classified the psychological impact of the outbreak as moderate or severe; 28.8% of respondents reported moderate to severe anxiety symptoms whereas 16.5% and 8.1% reported moderate to severe depressive symptoms and stress levels respectively(25). These findings are in line with the findings of the present study.

A recent study showed that 63.4% of victims with psychiatric disorders had comorbidity after Orissa super-cyclone in India (10) and another study showed that the prevalence of comorbid conditions was 14.3% between PTSD and depression, 24.9% between PTSD and anxiety and 13.1% between depression and anxiety in adolescents following earthquake in Indonesia (24). The prevalence of depression and anxiety comorbidity was found to be 23.2% (95% CI: 18.9-27.5) in this study, and this reasonably high rate of comorbidity justifies that the amount of damage to the general population caused by COVID-19 disease pandemic is similar to other emergencies. However, some differences in the prevalence of depression, anxiety and comorbid depression-anxiety might be due to the differences in tools used for assessing depression and anxiety and the study setting and study design.

In a previous study, the risk factors of depression and anxiety in survivors of an earthquake were reported to be age, pre and post-disaster traumatic incident, persisting violence, peri-traumatic distress, family and street violence (22). A study conducted among college students in South Korea found that female students (2.98) were more stressed than male students (2.84); also female students (0.66) had higher levels of anxiety compared to male students (0.50). Existing studies rigorously show that there were statistically significant differences in the risk of anxiety by sex, residence type, economic status, and Body Mass Index (BMI) (16). This is in line with the findings of the present study that females, compared to males, were 2.4 times (95% CI: 1.48-3.89) and 6.3 (95% CI: 3.54-11.18) more likely to have depression and anxiety.

One study found that women, who are often the main caregivers for injured, sick, elderly and family members with long-term disabilities, are worried about the future, and with husbands unemployed, and children out of school, they are mostly worried about feeding and taking care of their families (26). Another study reported that social, cultural and existing gender norms tend to make women relatively more vulnerable than men to mental health problems. Yet, women and girls have less access to or control over assets/resources such as information, education, health and wealth, which is necessary for the response to hazardous events (27). These might explain why women have a higher risk of depression and anxiety than men, and this is exacerbated especially due to the COVID-19 disease pandemic and the circumstances created by the lockdown in Nepal.

Accurate and up-to-date health information like treatment, local outbreak situation and precautionary measures (e.g., hand hygiene, wearing a mask) were associated with a lower psychological impact of the outbreak and lower levels of stress, anxiety, and depression (p < 0.05) (25). Likewise, another study reported that an individual with longer exposure to disaster-related news showed more symptoms of stress than those with less news exposure (16). This finding resembled the current study where the individuals who have higher exposure to media in accessing COVID-19 information have higher risk of depression, anxiety and comorbidity. Hence, emphasis should be given to provide correct and appropriate information on disaster events rather than describing the ravages caused by events, which could lead to an increase in unnecessary anxieties/fears related to transmission, testing positive, quarantine and stigma associated with COVID-19 disease, (16).

A prospective study conducted among a working-age population living alone and who used anti-depressant medications in Finland found that, during the 7-year follow-up, those who lived alone had an 80% higher risk of initiating antidepressant use compared to those who lived with family (28). Similarly, it was found that poor housing conditions were associated with increased use of antidepressants (28). In line with the findings of these studies, the present study also reported that living alone was found to be associated with depression (AOR: 3.5; 95% CI: 1.82-6.84) and anxiety (AOR: 3.3; 95% CI: 1.65-6.74) and the comorbid condition (AOR: 8.8, 95% CI: 3.85-20.12) compared to those who were living with their family during the lockdown period.

Likewise, participants with a health professional background had 1.7 times, 2 times and 3.4 times higher odds of having depression (95% CI: 1.04-2.90), anxiety (95% CI: 1.16-3.69) and depression-anxiety comorbidity (95% CI: 1.62-6.97) respectively. This finding resembled a meta-analysis which showed that one in three medical students have anxiety globally- a prevalence rate which is substantially higher than general population(29). This might be because health professionals perceive a higher level of risk associated with the COVID-19 disease, since they have easy access to information compared to the general population. Besides being fully aware of the risks, health professionals are known as the first responders to COVID-19 disease and thus, are more at risk for transmission.

## Conclusions

Depression, anxiety and depression and anxiety comorbidity are prevalent among the general population during the COVID-19 pandemic lockdown in Nepal. We identified the specific sub-groups of the general population at higher risk of depression, anxiety and the comorbid condition are females, those living alone during the COVID-19 pandemic lockdown, health professionals and those who spent more time accessing COVID-19 information. Governments should focus on disseminating appropriate knowledge about the disease using appropriate methods, and special interventions to promote the mental well-being need to be immediately implemented, with particular attention paid to high-risk groups. For instance, health workers are known to be at higher level of risk and thus should be prioritized when such interventions are implemented. Moreover, community mental health care should be made accessible to people who are at increased risk.

### Study Limitations

This study had several limitations that should be considered when interpreting the data. First, the use of an online survey imposes potential limitations. It is probable that the study findings under-represent the responses of those in certain demographics, e.g. those who are less educated, those less affluent, and elder age populations. Also, the online survey is relatively uncontrolled. The sample was self-selected and therefore there might be a high chance of response bias compared with a sample that had been randomly selected. Finally, this study does not rule out the association among COVID-19 lockdown and outcome variables but only assesses the prevalence and predicators during lockdown in Nepal.

## Data Availability

The data set for this study has been uploaded as the supplementary document.

## Acknowledgements

The authors would like to thank all the participants who participated in the survey. We would like to extend our sincere gratitude towards Ministry of Health and Population (MOHP), Nepal Public Health Association (NEPHA), Pokhara University and Purbanchal University for supporting in data collection.

## Author Contributions

Conceptualization: Anil Sigdel, Anu Bista, and Navaraj Bhattarai

Formal Analysis: Anil Sigdel and Navaraj Bhattari

Investigation: Bimal Chandra Pun, Govind Giri and Anu Bista

Methodology: Anil Sigdel, Bimal Chandra Pun, Anu Bista and Subash Thapa

Software: Anil Sigdel, Navaraj Bhattarai and Govind Giri

Writing-Original Draft Preparation: Anil Sigdel, Anu Bista, Subash Thapa and Hannah Marqusee

Writing-Reviewing & Editing: Anil Sigdel, Subash Thapa, Anu Bista and Hannah Marqusee

## Supporting Information

S1 Appendix. Questionnaire for the survey

## References

1. World Health Organization. 2019-nCoV outbreak is an emergency of international concern [Internet]. 2020 [cited 2020 Jan 4]. Available from: http://www.euro.who.int/en/health-topics/health-emergencies/international-health-regulations/news/news/2020/2/2019-ncov-outbreak-is-an-emergency-of-international-concern

2. Song P, Karako T. COVID-19: Real-time dissemination of scientific information to fight a public health emergency of international concern. Biosci Trends. 2020;14(1):1–2.

3. World Health Organizaton. COVID-19 CORONAVIRUS PANDEMIC [Internet]. 2020. [cited 2020 April 22]. Available from: https: https://covid19.who.int/

4. James H. Could coronavirus bring about the “waning of globalization”? [Internet]. World Economic Forum. 2020 [cited 2020 Jan 4]. Available from: https://www.weforum.org/agenda/2020/03/globalization-coronavirus-covid19-epidemic-change-economic-political

5. The Covid-19 outbreak so far and how Nepal can prepare for the worst. The Kathmandu Post [Internet]. 2020 Apr; Available from: https://kathmandupost.com/national/2020/03/21/the-covid-19-outbreak-so-far-and-how-nepal-can-prepare-for-the-worst

6. Coughlin SS. Anxiety and depression: linkages with viral diseases. Public Health Rev. 2012;34(2):1–17.

7. Makwana N. Disaster and its impact on mental health: A narrative review. J Fam Med Prim Care [Internet]. 2019;8(10):3090. Available from: http://www.jfmpc.com/article.asp?issn=2249-4863;year=2017;volume=6;issue=1;spage=169;epage=170;aulast=Faizi

8. World Health Organization. Mental Health in Emergencies [Internet]. 2019 [cited 2020 Jan 4]. Available from: https://www.who.int/news-room/fact-sheets/detail/mental-health-in-emergencies

9. Kar N. Indian research on disaster and mental health. Indian J Psychiatry. 2010;52(7):286.

10. Wachinger G, Renn O. Risk perception of natural hazards [Internet]. WP3-Report of the. 2010. Available from: http://caphaz-net.org/outcomes-results/CapHaz-Net_WP3_Risk-Perception2.pdf

11. World Health Organization. Depression and Others Common Mental Health Disorders: Global Health Estimates. Geneva; 2017.

12. World Health Organization. Depression Factsheet [Internet]. 2020 [cited 2020 Jan 4]. Available from: https://www.who.int/news-room/fact-sheets/detail/depression

13. Gong Y, Han T, Chen W, Dib HH, Yang G, Zhuang R, et al. Prevalence of anxiety and depressive symptoms and related risk factors among physicians in China: A cross-sectional study. PLoS One. 2014;9(7):1–7.

14. Bremmer I. Why COVID-19 May be a Major Blow to Globalization. TIME [Internet]. 2020 Mar; Available from: https://time.com/5796707/coronavirus-global-economy/

15. Nepal M of H and P. Coronavirus disease (COVID-19) outbreak updates & resource materials [Internet]. 2020 [cited 2020 April 22]. Available from: https://heoc.mohp.gov.np/recent_alert/update-on-novel-corona-virus-2019_ncov/

16. Lee E, Lee H. Disaster awareness and coping: Impact on stress, anxiety, and depression. Perspect Psychiatr Care. 2019;55(2):311–8.

17. Ministry of Health and Population (MoHP) NE and III. Nepal Demographic and Health Survey 2016. Kathmandu, Nepal; 2017.

18. Kroenke K, Spitzer RL, Williams JBW. The PHQ-9. J Gen Intern Med [Internet]. 2001 Sep;16(9):606–13. Available from: http://link.springer.com/10.1046/j.1525-1497.2001.016009606.x

19. Spitzer RL, Williams JBW, Kroenke K. Anxiety and PHQ-9 Depression score sheet [Internet]. 1999. Available from: https://www.torbayandsouthdevon.nhs.uk/uploads/score-sheet-gad-7-anxiety-and-phq-9-depression.pdf

20. Schaller M, Murray DR. Pathogens, Personality, and Culture: Disease Prevalence Predicts Worldwide Variability in Sociosexuality, Extraversion, and Openness to Experience. J Pers Soc Psychol. 2008;95(1):212–21.

21. Mortensen CR, Becker DV, Ackerman JM, Neuberg SL, Kenrick DT. Infection breeds reticence: The effects of disease salience on self-perceptions of personality and behavioral avoidance tendencies. Psychol Sci. 2010;21(3):440–7.

22. Derivois D, Cénat JM, Joseph NE, Karray A, Chahraoui K. Prevalence and determinants of post-traumatic stress disorder, anxiety and depression symptoms in street children survivors of the 2010 earthquake in Haiti, four years after. Child Abuse Negl [Internet]. 2017 May;67:174–81. Available from: https://linkinghub.elsevier.com/retrieve/pii/S0145213417300820

23. Maunder R, Hunter J, Vincent L, Bennett J, Peladeau N, Leszcz M, et al. The immediate psychological and occupational impact of the 2003 SARS outbreak in a teaching hospital. Cmaj. 2003;168(10):1245–51.

24. Marthoenis M, Ilyas A, Sofyan H, Schouler-Ocak M. Prevalence, comorbidity and predictors of post-traumatic stress disorder, depression, and anxiety in adolescents following an earthquake. Asian J Psychiatr [Internet]. 2019;43(May):154–9. Available from: https://doi.org/10.1016Zj.ajp.2019.05.030

25. Wang C, Pan R, Wan X, Tan Y, Xu L, Ho CS, et al. Immediate psychological responses and associated factors during the initial stage of the 2019 coronavirus disease (COVID-19) epidemic among the general population in China. Int J Environ Res Public Health. 2020;17(5).

26. Benites A&C. When disaster strikes: women’s particular vulnerabilities and amazing strengths. Vol. 53, Trabajo Infantil. 2016.

27. Network D. Women, Girls and Disasters A review for DFID by Sarah Bradshaw 1 and Maureen Fordham 2. 2013.

28. Pulkki-råback L, Kivimäki M, Ahola K, Joutsenniemi K, Elovainio M, Rossi H, et al. Living alone and antidepressant medication use: a prospective study in a working-age population. BMC Public Health [Internet]. 2012;12(1):236. Available from: http://www.biomedcentral.com/1471-2458/12/236

29. Quek TTC, Tam WWS, Tran BX, Zhang M, Zhang Z, Ho CSH, et al. The global prevalence of anxiety among medical students: A meta-analysis. Int J Environ Res Public Health. 2019;16(15).

